# Factors associated with COVID-19 vaccine acceptance and hesitancy among residents of Northern California jails

**DOI:** 10.1101/2021.11.22.21266559

**Authors:** Yiran E Liu, Jillian Oto, John Will, Christopher LeBoa, Alexis Doyle, Neil Rens, Shelley Aggarwal, Iryna Kalish, Marcela Rodriguez, Beruk Sherif, Chrisele Trinidad, Michael Del Rosario, Sophie Allen, Robert Spencer, Carlos Morales, Alexander Chyorny, Jason R Andrews

**Affiliations:** Department of Medicine, Division of Infectious Diseases and Geographic Medicine, Stanford University School of Medicine, Stanford, California, USA; Cancer Biology Graduate Program, Stanford University School of Medicine, Stanford, California, USA; Custody Health Services, Santa Clara Valley Health and Hospital System, San Jose, California, USA; Stanford University School of Medicine, Stanford, California, USA; Department of Pediatrics, Santa Clara Valley Health and Hospital System, San Jose, California, USA; Department of Medicine, Division of Custody Health, Santa Clara Valley Health and Hospital System, San Jose, California, USA; Stanford Center for Clinical Research, Stanford University School of Medicine, Stanford, California, USA; Division of Correctional Health Services, San Mateo County Health, Redwood City, California, USA; Stanford Law School, Stanford, California, USA; Department of Sociology, Stanford School of Humanities and Sciences, Stanford, California, USA

**Author notes:** these authors contributed equally (Yiran E Liu, Jillian Oto, John Will). these authors contributed equally (Alexander Chyorny, Jason R Andrews).

## Abstract

**Background:** Carceral facilities are high-risk settings for COVID-19 transmission. Understanding of factors associated with COVID-19 vaccine acceptance and hesitancy among incarcerated individuals is incomplete, especially for people living in jails.

**Methods:** We conducted a retrospective review of COVID-19 vaccination data from the electronic health record (EHR) of residents in two Northern California county jails to examine factors associated with vaccine uptake in this population. We additionally administered a survey in four jails to assess reasons for vaccine hesitancy, sources of COVID-19 information, and medical mistrust. We performed multivariate logistic regression to determine associations with vaccine uptake or hesitancy.

**Results:** Of 2,584 jail residents offered a COVID-19 vaccine between March 19, 2021 and June 30, 2021, 1,464 (56.7%) accepted at least one dose. Among vaccinated residents, 538 (36.7%) initially refused the vaccine. Vaccine uptake was higher among older individuals, women, those with recent flu vaccination, and those living in shared cells or open dorms. Leading reasons for vaccine hesitancy included concerns around side effects and suboptimal efficacy. Television and friends/family were the most commonly cited and the most trusted sources of COVID-19 information, respectively. Vaccine acceptance was associated with increased trust in COVID-19 information sources and in medical personnel both in and out of jail.

**Conclusion:** Ongoing evidence-based COVID-19 vaccination efforts are needed in high-risk carceral settings. Effective interventions to improve vaccination rates in this population should utilize accessible and trusted sources of information to address concerns about vaccine side effects and efficacy and foster medical trust.

## INTRODUCTION

Prisons, jails, and detention centers have been dangerous settings for COVID-19 transmission [1, 2], with some of the largest outbreaks in the US to-date [3]. Moreover, policing and incarceration disproportionately affect individuals and communities that already experience increased risk of COVID-19 infection and death due to systemic inequities, medical comorbidities, substance use disorders, mental health illnesses, and housing insecurity [4-8].

High vaccine coverage is needed to mitigate COVID-19 transmission in congregate, high-density settings such as prisons and jails, especially in the context of existing and future viral variants [9, 10]. Despite the importance of achieving high vaccination rates in this population, there is a paucity of data on COVID-19 vaccine acceptance and reasons for hesitancy among incarcerated individuals to guide evidence-based vaccination efforts. Recent studies in US prisons have identified differences in COVID-19 vaccine acceptance by age, gender/sex, and race/ethnicity [11, 12]. However, less is known about COVID-19 vaccination among people living in jails, where there is more frequent turnover than in prisons and where many people have not been convicted of a crime [13]. Surveys conducted prior to vaccine availability have suggested lower willingness to get a COVID-19 vaccine among jail residents than prison residents [14, 15], but to our knowledge there have been no studies on actual COVID-19 vaccine uptake in jails. Moreover, while it is hypothesized that incarcerated individuals may harbor mistrust of medical, custodial, or governmental personnel or institutions [16], the association of this mistrust with COVID-19 vaccine acceptance among incarcerated individuals is poorly understood.

In this study, we utilized electronic health record (EHR) data of COVID-19 vaccination in two Northern California jails and investigated the association between COVID-19 vaccination and age, gender, race/ethnicity, recent flu vaccination, and housing type. We additionally conducted a survey among incarcerated individuals across four jails to examine potential factors underlying vaccine hesitancy, including mistrust of medical personnel and sources of COVID-19 information.

## METHODS

### Jail Vaccination Program

In Santa Clara County and San Mateo County, COVID-19 vaccines became available to older and medically vulnerable jail residents on January 29, 2021 and to all jail residents on March 19, 2021. Residents were screened periodically for interest in COVID-19 vaccination; anyone who expressed interest during screening was subsequently scheduled, upon becoming eligible, for an upcoming on-site vaccination clinic. Vaccination clinics occurred biweekly in individual housing units. Individuals could change their decision regarding vaccination at any time, including during a regular medical appointment, when called for vaccination, or via the typical medical request process. In Santa Clara County, all data related to vaccination in custody, including each instance of screening and refusals, were entered in the electronic health record (EHR). In San Mateo County, only data on vaccinations, but not refusals, were recorded by Correctional Health Services.

### Retrospective Chart Review and Analysis of Factors Associated with Vaccine Uptake

We searched the Santa Clara County EHR for individuals screened and/or vaccinated in custody between 1/29/2021 and 6/30/2021. We limited the observation period for vaccine uptake to individuals admitted on or after 3/19/2021 with a minimum incarceration length of 26 days (the longest time between vaccine clinics at any individual jail). We excluded those who received their first dose outside of jail, prior to admission.

We performed multivariable logistic regression to determine the association between COVID-19 vaccine uptake and age, gender, race/ethnicity, recent flu vaccination (within the last two years, according to the EHR and state vaccination repository), and housing type. Housing type (single cell, shared/double cell, open dorm) was defined by the unit in which an individual resided at the time of vaccination, or for those unvaccinated, at the time of the most recent screening. Open dorms are defined as units where there are at least three individuals housed together and typically consist of between 40 and 80 people.

### Survey on Vaccine Attitudes, Sources of COVID-19 Information, and Medical Trust

We additionally conducted a survey to assess attitudes toward vaccination among jail residents in both counties. The survey was part of an ongoing study on perceptions, attitudes, and behaviors surrounding COVID-19 in these jails (Stanford IRB 56169, Valley Medical Center IRB 20-022). Between July 8, 2020 and April 30, 2021 we enrolled 788 incarcerated participants. Participants were recruited through flyers and announcements in their housing unit; in single-cell units, recruitment was done door-to-door. After administering written informed consent, surveys were administered by research coordinators via an electronic tablet; participants could choose to read and respond to questions themselves or respond orally to questions read aloud. Spanish translations were available. Survey data were recorded in a HIPAA-secure REDCap database [17].

Beginning December 15, 2020, we added survey questions about vaccination and medical trust (**S1 Survey**), after which 509 participants responded. Questions on vaccine acceptance and hesitancy were adapted from national surveys conducted by the Pew research group [18], and questions on medical trust were adapted from prior research on the relationship between medical trust and patient behavior in Oakland, California [19]. We linked participants’ survey responses to records of their vaccine uptake through June 30, 2021 (**S2 Text**). For 90 respondents, data were missing or insufficient to determine vaccine refusal or uptake, likely due to release from jail prior to vaccine eligibility.

For analysis of survey responses, we defined vaccine acceptance as intent to get a COVID-19 vaccine. For 125 respondents who had already been vaccinated or had already refused prior to participating in the survey, we defined vaccine acceptance as uptake or refusal. We performed chi-square tests of independence to determine whether differences in vaccine acceptance among subgroups were statistically significant. Respondents who selected “Prefer not to answer” for a given question were excluded from analysis of that question.

### Analysis of Vaccine Acceptance and Medical Trust

We conducted multivariable logistic regression to investigate the effect of medical trust on vaccine acceptance among survey respondents. Predictor variables included age group (18-29, 30-49, 50+), gender (man, woman, transgender person), race/ethnicity (Hispanic/Latinx, non-Hispanic Black, non-Hispanic White, non-Hispanic Asian, non-Hispanic other/unknown race), general trust (“Generally speaking, would you say that most people can be trusted?”), medical trust (trust, neutral/do not trust), and the interaction between race/ethnicity and medical trust. Separate regressions were fit using either trust in jail health staff or trust in one’s doctor as medical trust. Trust in jail health staff was assessed using the question, “I trust the jail doctors’ and nurses’ judgments about my medical care.” Trust in one’s outside doctor was assessed using the question, “I trust my doctor’s judgments about my medical care.” We computed adjusted odds ratios for the difference in vaccine acceptance between those within each racial/ethnic group who trust jail health staff/their doctor versus those who are neutral or do not trust jail health staff/their doctor.

Analyses were performed using Stata version 17.0 and R version 4.0.5.

## RESULTS

### COVID-19 vaccination among jail residents varied by demographic factors, housing unit type, and recent flu vaccination

Between March 19, 2021 and June 30, 2021, 2,584 jail residents were offered a COVID-19 vaccine while incarcerated, of whom 1,464 (56.7%) accepted at least one dose. Results of a multivariable logistic regression showed that acceptance increased with age and that women were significantly more likely to get a COVID-19 vaccine (**Table 1**). Vaccine uptake was lowest among Black individuals (47.7%) but otherwise did not vary substantially by race/ethnicity. Those with a recent flu vaccination were 2.7 (95% CI, 2.2-3.4) times more likely to get a COVID-19 vaccine in custody. Individuals housed in shared cells and open dorms were significantly more likely to get vaccinated than those housed in private cells.

**Table 1.** Vaccination and adjusted odds ratios for vaccine uptake by demographic characteristics, recent flu vaccination, and housing type.

| Group | Number Offered Vaccine (% of total) | Number Vaccinated (% of subgroup) | Adjusted Odds Ratio (95% CI) |
| --- | --- | --- | --- |
| **Age** |  |  |  |
| 18-29 | 778 (30.1) | 324 (41.7) | Ref |
| 30-49 | 1,468 (56.8) | 897 (61.1) | 2.2 (1.8-2.6)*** |
| 50+ | 338 (13.1) | 243 (71.9) | 3.5 (2.6-4.6)*** |
| **Gender** |  |  |  |
| Female | 258 (10.0) | 163 (63.2) | Ref |
| Male | 2,326 (90.0) | 1,301 (55.9) | .6 (.5-.8)*** |
| **Race/ethnicity** |  |  |  |
| White | 476 (18.4) | 275 (57.8) | Ref |
| Hispanic/Latinx | 1,405 (54.4) | 809 (57.6) | 1.1 (0.9-1.4) |
| Black | 298 (11.5) | 142 (47.7) | 0.7 (0.5-1.0) |
| Asian | 206 (8.0) | 126 (61.2) | 1.2 (0.8-1.6) |
| Other/Unknown | 199 (7.7) | 112 (56.3) | 1.1 (0.8-1.6) |
| **Recent Flu Vaccination** |  |  |  |
| No / Unknown | 2,041 (79.0) | 1,058 (51.8) | Ref |
| Yes | 543 (21.0) | 408 (75.1) | 2.7 (2.2-3.4)*** |
| **Housing Type** |  |  |  |
| Single Cell | 149 (5.8) | 62 (41.6) | Ref |
| Shared Cell | 1,163 (45.0) | 616 (53.0) | 1.8 (1.2-2.6)** |
| Open Dorm | 1,272 (49.2) | 786 (61.8) | 2.3 (1.6-3.4)*** |
| **Total** | 2,584 | 1,464 (56.7) |  |

\*\*, $p \leq 0.01$ ; \*\*\*, $p \leq 0.001$ . Recent flu vaccination was defined as documentation within the last two years in the EHR and state vaccination repository.

### Individual-level changes in vaccine acceptance

In an attempt to increase vaccination rates, multiple offers for vaccination were made to those in custody who had previously declined the vaccine. Of the 1,464 individuals vaccinated in jail, 538 (36.7%) had indicated during at least one previous screening that they did not wish to get the COVID-19 vaccine. Those who were ultimately vaccinated despite previous refusal (i.e. who changed their minds from “No” to “Yes”), were more likely to be younger, women, and residing in shared cells and open dorms (**Fig S1A)**. Conversely, of the 1,120 individuals who ultimately did not get vaccinated, 161 (14.4%) had expressed interest in receiving the COVID-19 vaccine during a previous screening. Among those who remained unvaccinated, the percentage who had previously expressed interest (“Yes to No”) ranged from 11% to 17% across demographic groups (**Fig S1B)**.

### Reasons for COVID-19 vaccine hesitancy

Between December 15, 2020 and April 30, 2021 we surveyed 509 jail residents about COVID-19 vaccination, sources of information, and medical trust (**Table S1)**. The leading reasons for hesitancy among survey respondents who did not intend to get a COVID-19 vaccine or who refused the vaccine (N=140, “vaccine-hesitant respondents”) were concern about side effects (60%) and wanting to know more about how well it works (38%), with most citing these as major reasons (**Fig 1**). Twenty-three percent of vaccine-hesitant respondents indicated that they did not think they needed it.

**Figure 1.**
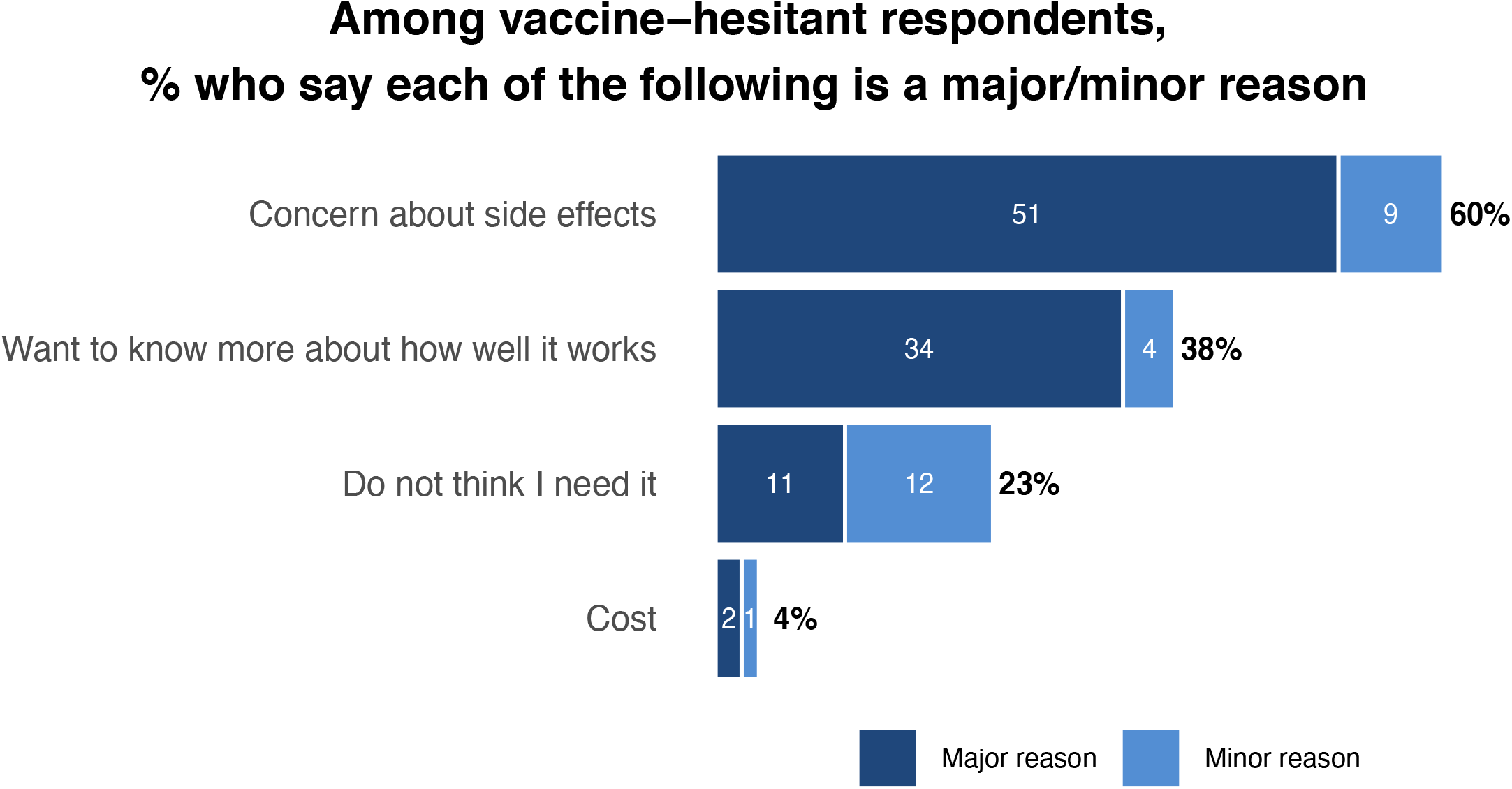
Reasons for vaccine hesitancy among survey respondents who did not intend to get a COVID-19 vaccine or who already refused the vaccine. Proportion of vaccine-hesitant participants who report each as a major (dark blue) or minor (light blue) reason. White numbers indicate percentage of vaccine-hesitant respondents who selected each as a major/minor reason, black percentages in bold indicate total percentage of vaccine-hesitant respondents who selected each reason.

To examine potential factors underlying individual-level changes in vaccine acceptance, we compared deterrents to vaccination among survey respondents with consistent or discordant intent and uptake. Survey participants who were initially hesitant but eventually got vaccinated (“No to Yes”) were more likely to cite suboptimal efficacy and cost than hesitant individuals that did not get vaccinated (“Consistent No”). They were also less likely to cite side effects and annual boosters as factors influencing their decision than the “Consistent No” group (**Fig S2**). Conversely, individuals who intended to get vaccinated but ultimately did not (“Yes to No”) were more likely than individuals who had consistent intent and uptake (“Consistent Yes”) to cite all factors as deterrents to vaccination.

### Medical mistrust

When asked about their doctor outside of jail, 65.8% of survey participants reported trusting their doctor’s judgments about their medical care, and 24.9% and 9.3% said they were neutral toward or did not trust their doctor’s judgments, respectively (**Fig S3**). Trust in jail health staff was much lower, with 35.8%, 34.7%, and 29.4% of respondents who said they felt trustful, neutral, or mistrustful, respectively, toward jail health staff’s judgments about their medical care. Medical trust, both for one’s outside doctor and jail health staff, was highest among white respondents compared to Latinx or Black respondents.

Trust in jail health staff was associated with COVID-19 vaccine acceptance (p<0.001), with 81%, 72%, and 59% acceptance among respondents who reported feeling trustful, neutral, or distrustful, respectively, toward jail health staff (**Fig. 2A**). However, this association was not generalizable across racial/ethnic groups. After adjusting for age, gender, and general trust in others, our results showed that trust in jail health staff was associated with 2.8 (95% CI, 0.7-11.6) and 2.5 (95% CI, 1.2-5.1) times greater vaccine acceptance among white and Latinx participants, respectively, but had no effect on COVID-19 vaccine acceptance among participants of other races (**Fig. 2B**). Similar trends were found with trust in one’s outside doctor (**Fig S4**).

**Figure 2.**
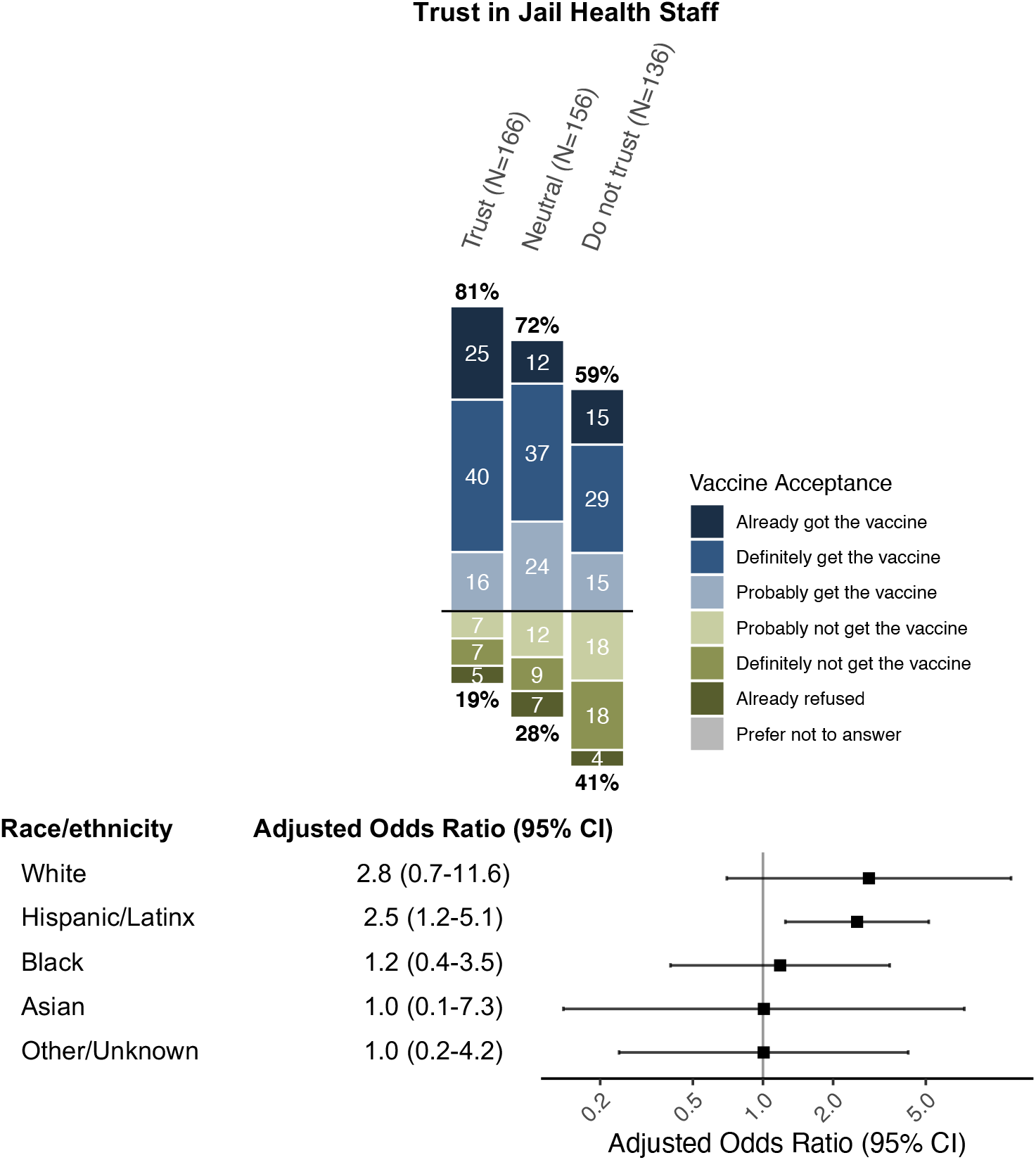
Association between trust in jail health staff and COVID-19 vaccine acceptance among survey respondents. A) Proportion of respondents, stratified by trust in jail health staff, who indicated vaccine acceptance (above the horizontal black line) or hesitancy (below line). White numbers indicate percentages within each vaccine acceptance level; black percentages in bold indicate total percentages of vaccine-accepting (top) or vaccine-hesitant (bottom) respondents in each trust strata. The difference in vaccine acceptance/hesitancy among different trust strata was statistically significant by the chi-square test of independence, with p-value < 0.001. B) Results of a multivariate logistic regression adjusted for age, gender, and general trust in others. The adjusted odds ratio reflects the increase in likelihood of vaccine acceptance among respondents in each racial/ethnic group who trust jail health staff. Respondents who selected “Prefer not to answer” for the vaccine and/or trust question (N=51) were excluded from this analysis.

### Vaccine acceptance and sources of COVID-19 information

Television was the leading source of information about COVID-19 cited by 65% of survey respondents (**Fig 3**). Friends and family were the next most common (43%), and most trusted, source of information. Reported sources of information were not considerably different by vaccine acceptance; however, those who already got or intended to get a vaccine were more likely to trust most sources of information (**Fig S5**).

**Figure 3.**
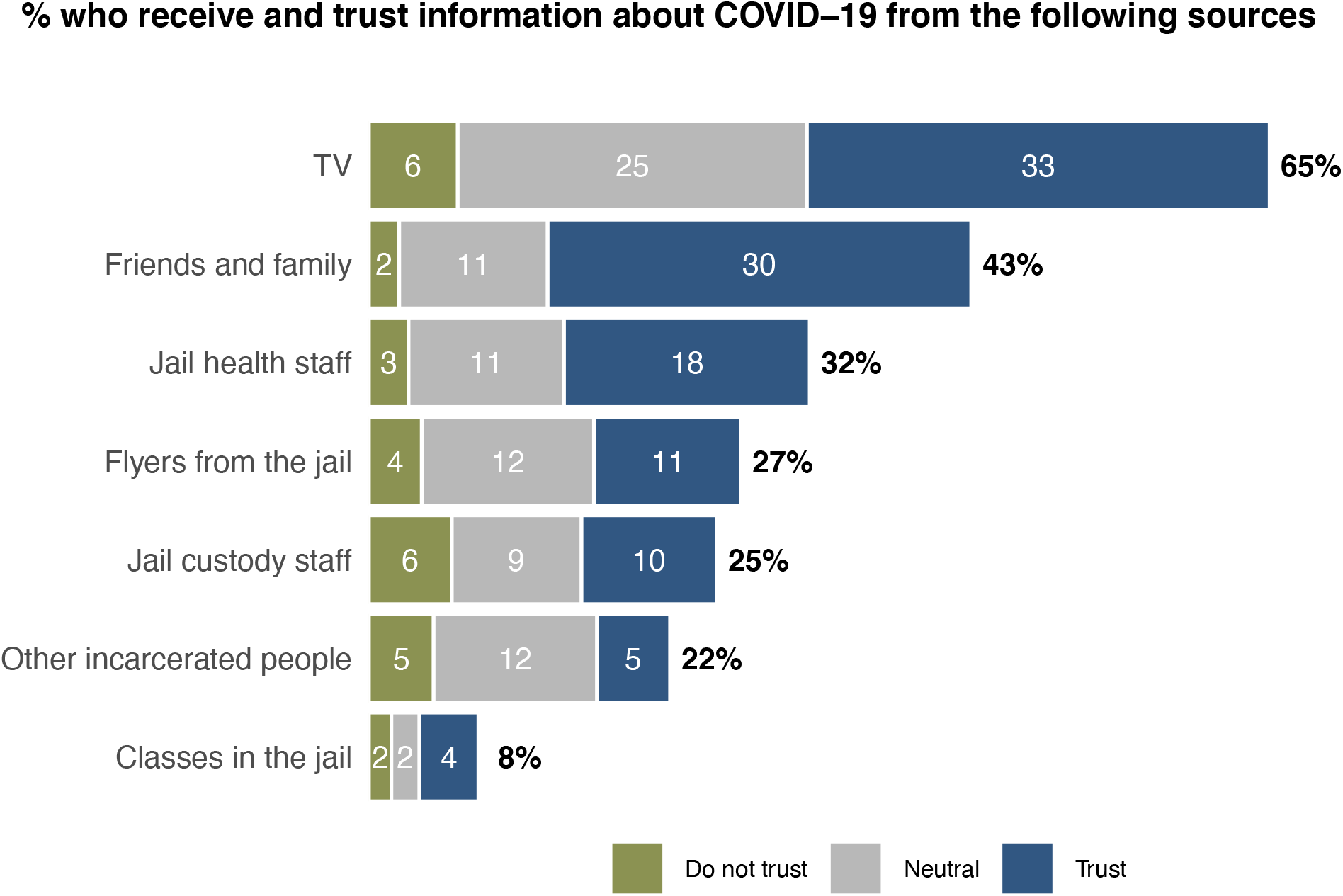
Sources of information about COVID-19 and trust in each source among survey respondents. Proportion of respondents who report receiving information about COVID-19 from each source (black percentages in bold), and who trust, do not trust, or are neutral toward each source (white numbers).

## DISCUSSION

Vaccination is one strategy to reduce COVID-19 morbidity and mortality in high-risk settings like prisons and jails. However, our understanding of the factors underlying vaccine acceptance and hesitancy among incarcerated individuals is incomplete, particularly for those residing in jails. By reviewing the EHR of individuals incarcerated in two Northern California jails, we found that older age, female gender, recent flu vaccination, and shared housing were associated with increased COVID-19 vaccine uptake. Among those who were ultimately vaccinated, over one-third had previously declined; younger individuals, women, and those in shared housing were more likely than their counterparts to have gotten vaccinated after a prior refusal. A survey conducted across four jails revealed that concerns about side effects and suboptimal efficacy were leading reasons for vaccine hesitancy. Moreover, vaccine acceptance was associated with trust in COVID-19 information sources and in medical personnel, although the latter association varied substantially by race/ethnicity.

Consistent with previous studies on incarcerated and non-incarcerated populations [11, 12, 20], we found higher COVID-19 vaccine uptake among older jail residents. However, while a recent report in US prisons showed lower COVID-19 vaccine uptake among incarcerated women than men [12], we found the opposite, perhaps due to differences in setting, population, period of observation, and/or distinct policies affecting COVID-19 risk and housing unit culture for women in these jails. Existing research also shows gender disparities in healthcare utilization, with women utilizing services and preventative health measures more frequently than men [21]. Interestingly, nearly half of women who ultimately got the vaccine had previously declined, as was the case with individuals aged 18-29, suggesting that certain groups exhibiting hesitancy may be more likely to change their minds. In fact, across all groups, at least a quarter of those who had a history of a screening refusal eventually got vaccinated, demonstrating the importance of continued offers and availability. Conversely, of those who did not get vaccinated, 14.4% did show interest during a prior screening, suggesting that uptake may increase if vaccines were readily available at the time of screening.

Compared to people housed in single cell units, those in shared cells and open dorm settings were more likely to get vaccinated--both overall and after initial refusal. This finding may be partially confounded by differences in population characteristics in each housing type but may reflect possible peer influence and/or greater willingness to get vaccinated among those who witnessed firsthand the major outbreaks in shared cell and open dorm units in late 2020 through early 2021. Not surprisingly, individuals with a history of recent flu vaccination were more likely to get vaccinated for COVID-19, confirming prior surveys [22, 23] and perhaps reflecting more familiarity with and trust in the vaccination process in general.

Survey results showed that the most cited reasons for vaccine hesitancy surrounded side effects and efficacy, reflecting concerns reported in other studies [15, 18]. Additional research could be conducted to determine whether FDA approval of the COVID-19 vaccine(s) would have increased vaccine acceptance [24]. Approximately one third of already vaccine-hesitant respondents said they would be further deterred from getting a vaccine if an annual booster were needed, a finding with important implications as more data on long-term efficacy and booster recommendations emerge.

Although COVID-19 vaccines are currently free of charge in and out of custody, 4% of respondents nonetheless cited this as a reason for hesitancy; further, many respondents indicated that having to pay would make them less likely to get a vaccine. These results highlight the need for the continuation of, and clearer information about, free access to vaccines. Importantly, our findings call for such information to be disseminated through accessible, varied, and trusted sources as identified by justice-involved populations. In the present study, survey respondents, including those who were vaccine-hesitant, cited family and friends as the most trusted source of COVID-19 information. Of note, contact with loved ones declined during the pandemic due to suspension of in-person visitation. Nonetheless, this finding suggests that vaccine information campaigns outside of jail may also indirectly reach and influence vaccine attitudes among incarcerated individuals.

This study provides mixed support for the relationship between medical trust and vaccine acceptance among incarcerated individuals. Trust in jail health staff was lower than trust in one’s doctor outside of jail, but both were associated with higher vaccine acceptance overall. Notably, this association was absent among Black respondents, a surprising result given the frequently evoked link between lower COVID-19 vaccination rates and medical mistrust among Black Americans [25-27]. However, our sample size of Black respondents was small (N=77), and additional research is needed to validate this finding as well as to identify more salient and specific reasons for vaccine hesitancy among Black incarcerated individuals.

This study has several limitations. First, our retrospective chart review was restricted to data on vaccination in custody and did not include data on vaccination in the community following release. The chart review also excluded individuals who were in custody for under 26 days; future research should examine vaccine acceptance among those who are frequently admitted and released from custody as they may represent a uniquely vulnerable group and may facilitate COVID-19 transmission between jails and the community [28]. Next, the survey findings were limited by sample size and may not be representative of the entire jail population; they are also subject to limitations of self-reported data. Furthermore, although both chart review and survey enrollment occurred across several months, we did not have sufficient power to examine effects of calendar time on vaccine acceptance, as shown elsewhere [23]. While we attempted to infer factors underlying hesitancy and acceptance by linking survey responses with vaccination data, our understanding would be enhanced by more open-ended questions and qualitative data. Finally, these findings may have limited generalizability to the non-incarcerated population as well as to other carceral settings, as jails are often unique in culture, environment, and population.

## Conclusion

This study provides insight into the factors associated with COVID-19 vaccine acceptance among jail residents and identifies reasons for vaccine hesitancy and the most common and trusted sources of COVID-19 information in this population. More research is needed to inform evidence-based vaccination campaigns in this population, as incarcerated individuals continue to face increased risk of infection and mortality from COVID-19 [2] and other vaccine-preventable infectious diseases [29]. Furthermore, given the disproportionate incarceration of marginalized groups who have borne the brunt of COVID-19, efforts to increase vaccination in this population may also reduce COVID-19 disparities at large.

It bears noting that although custody health departments have an opportunity to provide health services like vaccinations for individuals who may face undue barriers in the community, they may struggle to achieve adequate uptake in part due to prevalent mistrust among their patients, as illustrated by our findings. Additional research may shed light on ways to improve trust in custody health providers, which may in turn improve vaccination rates among jail residents. Nonetheless, given the high coverage needed in congregate settings and amidst new viral variants, efforts to increase vaccination rates should be carried out alongside other measures to reduce transmission, including face masks, regular testing, and decarceration [30]. Together, these measures can mitigate COVID-19 transmission among vulnerable populations, reduce COVID-19 disparities, and improve population health.

## Supporting information

S1 Survey

STROBE Checklist

## Data Availability

Data from the present study are not currently available due to ethical and privacy concerns but may be made available in de-identified or aggregate form in the future, as allowed by the IRB.

## SUPPLEMENTARY MATERIALS

**Table S1. Characteristics of survey respondents**. Percentages may not sum up to 100 due to rounding. We define “already refused” as those who had a record of refusal prior to taking the survey and who still did not have a vaccination record by the end of the observation period (June 30, 2021).

**Figure S1. Individual-level changes in decision to get a COVID-19 vaccine among jail residents**. A) Proportion of ultimately vaccinated individuals who had a history of prior refusal (light blue, “No to Yes”), stratified by age, gender, housing, and race/ethnicity. B) Proportion of ultimately unvaccinated individuals who had indicated interest in a prior screening (light red, “Yes to No”), stratified by age, gender, housing, and race/ethnicity.

**Figure S2. Potential deterrents to vaccination, stratified by individual-level consistency or reversal in decision to get a COVID-19 vaccine**. Proportion of survey respondents in each group who say each of the following factors would make them a lot (dark blue) or a little (light blue) less likely to get a vaccine. “Consistent No” refers to respondents who did not intend to get a vaccine and who remained unvaccinated by the end of the observation period. “No to Yes” refers to those who did not intend to get a vaccine but who ultimately got vaccinated. “Consistent Yes” refers to those who intended to and ultimately got a vaccine. “Yes to No” refers to those who intended to get a vaccine but remained unvaccinated by the end of the study period.

**Figure S3. Trust in one’s outside doctor and in jail health staff among survey respondents**. Proportion of A) all respondents or B) respondents stratified by race/ethnicity who said they felt trustful, neutral, or distrustful toward their doctor outside of jail. Proportion of C) all respondents or D) respondents stratified by race/ethnicity who said they felt trustful, neutral, or distrustful toward jail health staff. Respondents who selected “Prefer not to answer” (N=36 and 37 for trust in outside doctor and trust in jail health staff, respectively) were excluded from this analysis.

**Figure S4. Association between trust in one’s outside doctor and COVID-19 vaccine acceptance among survey respondents**. A) Proportion of respondents, stratified by trust in one’s doctor outside of jail, who indicated vaccine acceptance (above the horizontal black line) or hesitancy (below line). White numbers indicate percentages within each vaccine acceptance level; black percentages in bold indicate total percentages of vaccine-accepting (top) or vaccine-hesitant (bottom) respondents in each trust strata. The difference in vaccine acceptance/hesitancy among different trust strata was statistically significant by the chi-square test of independence, with p-value < 0.001. B) Results of a multivariate logistic regression adjusted for age, gender, and general trust in others. The adjusted odds ratio reflects the increase in likelihood of vaccine acceptance among respondents in each racial/ethnic group who trust one’s doctor outside of jail. Respondents who selected “Prefer not to answer” for the vaccine and/or trust question (N=52) were excluded from this analysis.

**Figure S5. Sources of information about COVID-19 and trust in each source among survey respondents, stratified by vaccine acceptance**. Proportion of respondents, stratified by acceptance (those who would get or already got the vaccine) and hesitancy (those who would NOT get or already refused the vaccine), who report receiving information about COVID-19 from each source (black percentages in bold), and who trust, do not trust, or are neutral toward each source (white numbers).

**S1 Survey**. Survey questions relating to vaccination and medical trust.

**S2 Text**. Methods for linkage of survey data with vaccine uptake data.

## Licence statement

I, the Submitting Author has the right to grant and does grant on behalf of all authors of the Work (as defined in the below author licence), an exclusive licence and/or a non-exclusive licence for contributions from authors who are: i) UK Crown employees; ii) where BMJ has agreed a CC-BY licence shall apply, and/or iii) in accordance with the terms applicable for US Federal Government officers or employees acting as part of their official duties; on a worldwide, perpetual, irrevocable, royalty-free basis to BMJ Publishing Group Ltd (“BMJ”) its licensees and where the relevant Journal is co-owned by BMJ to the co-owners of the Journal, to publish the Work in Journal of Epidemiology & Community Health and any other BMJ products and to exploit all rights, as set out in our <u>licence</u>.

