## Supplementary figures and images for "Factors associated with COVID-19 vaccine acceptance and hesitancy among residents of Northern California jails"

### Supplementary Figures

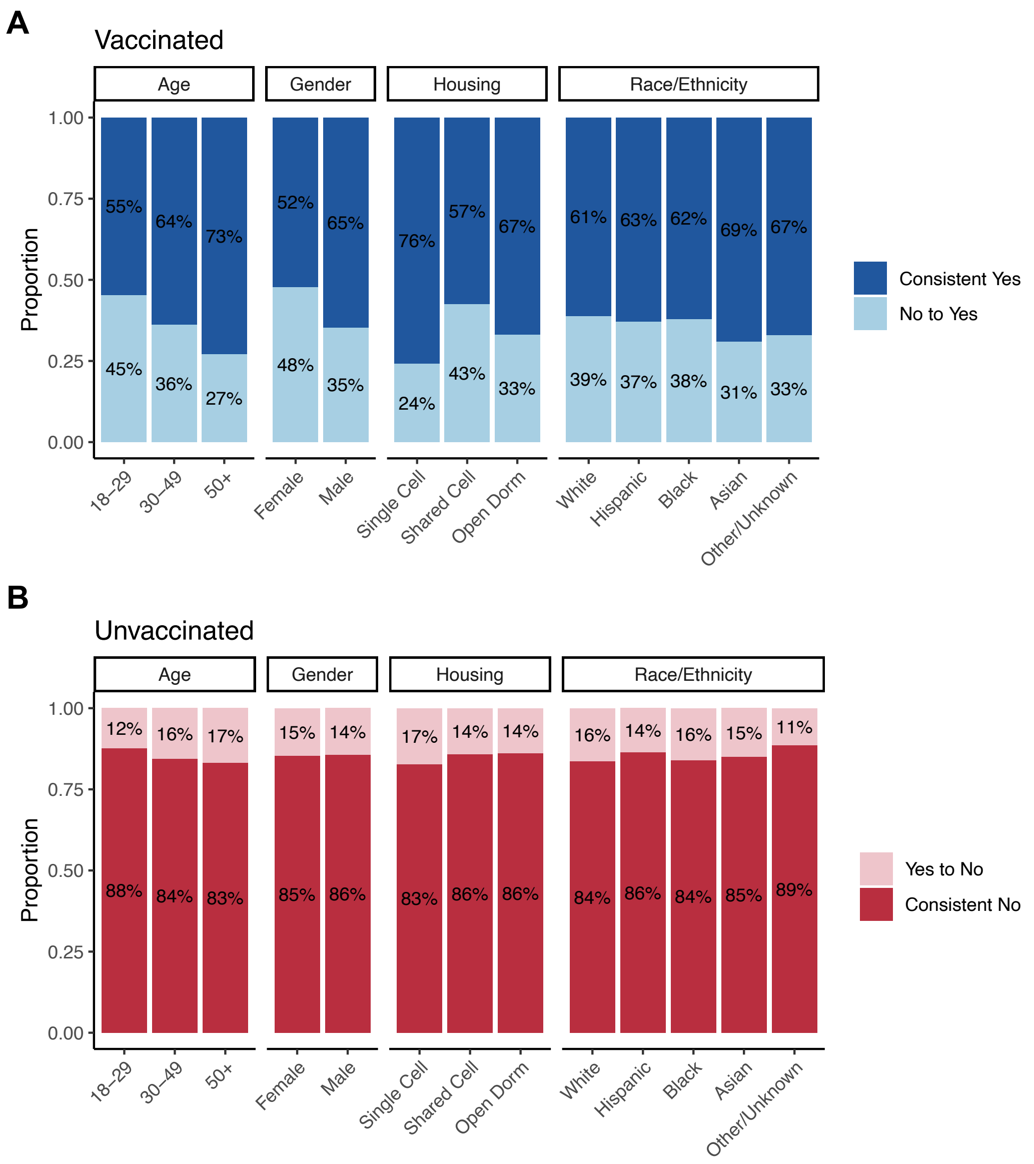

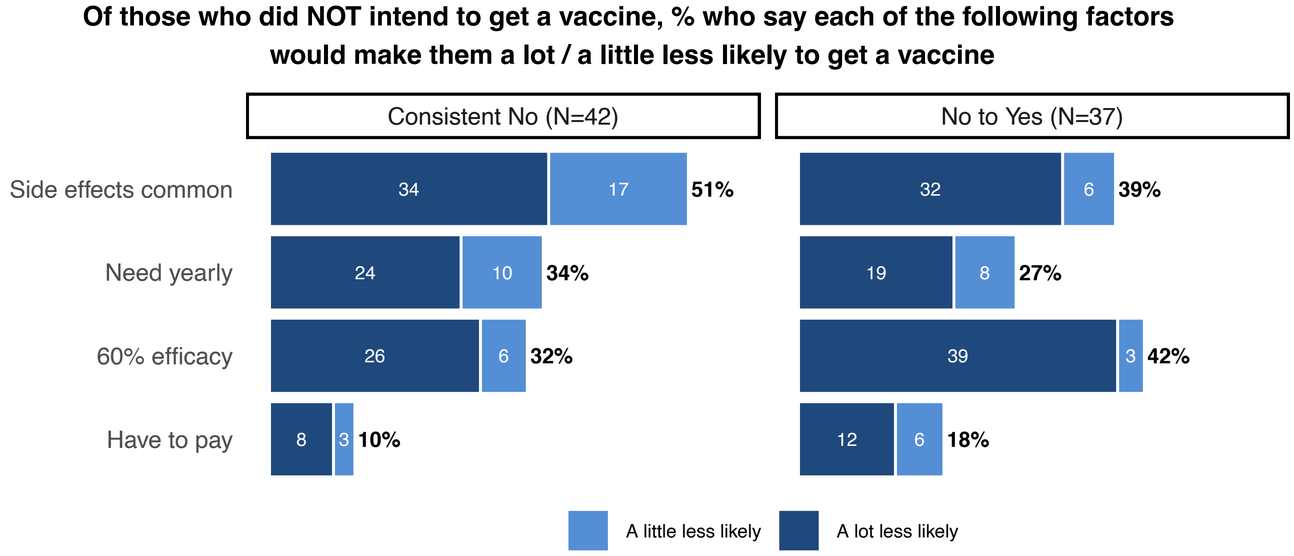

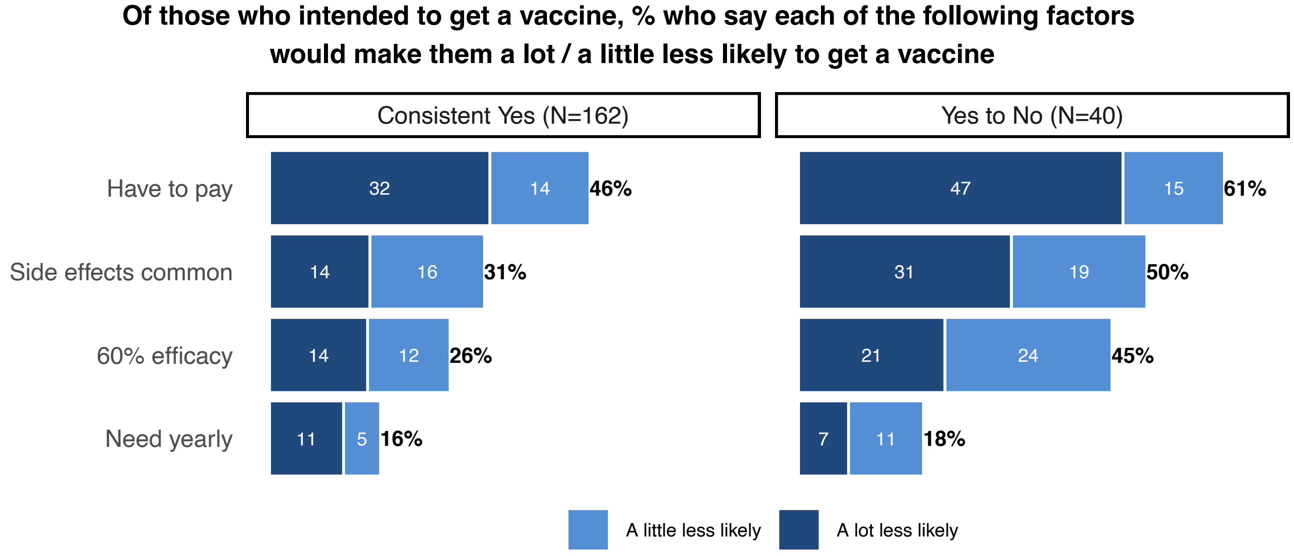

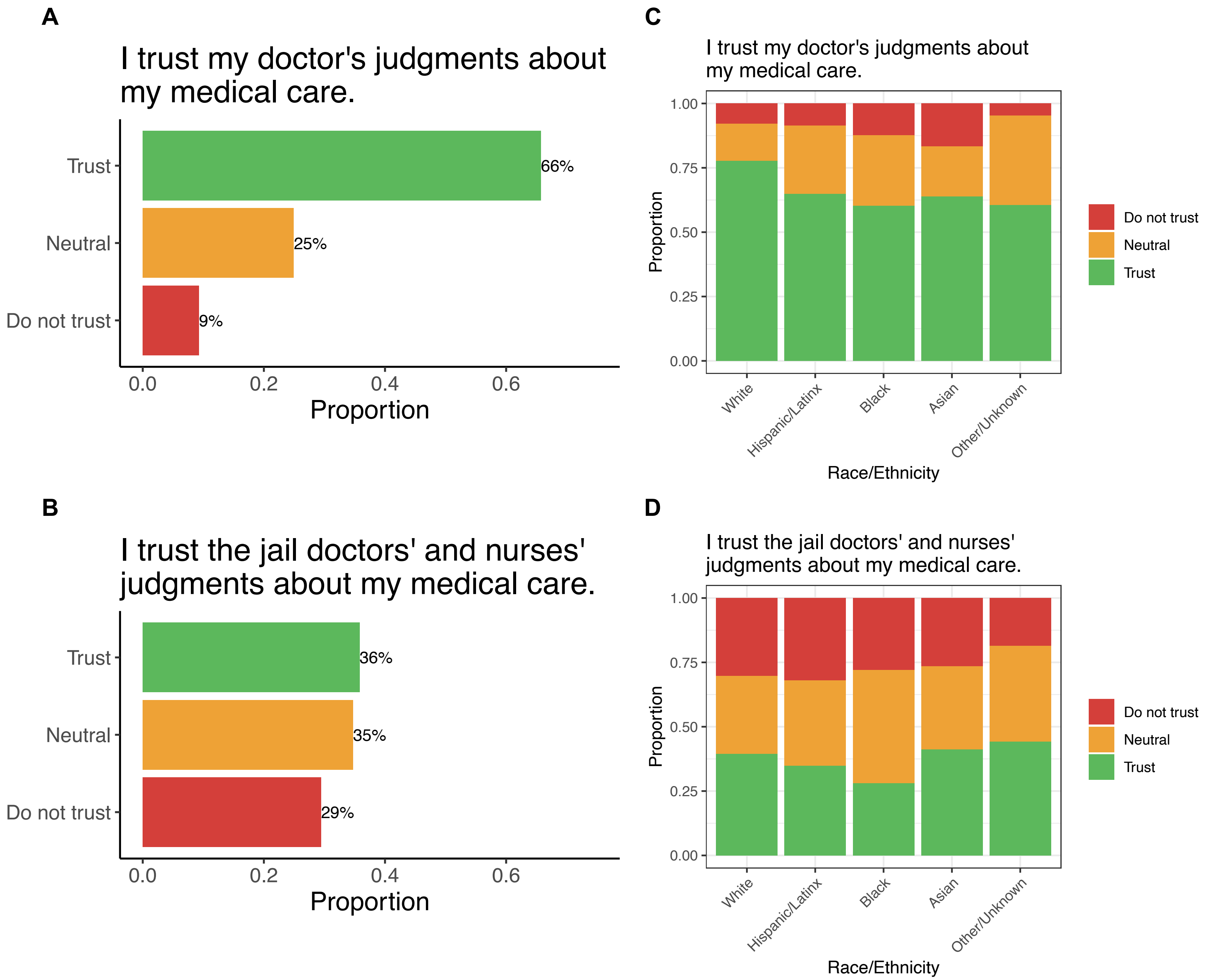

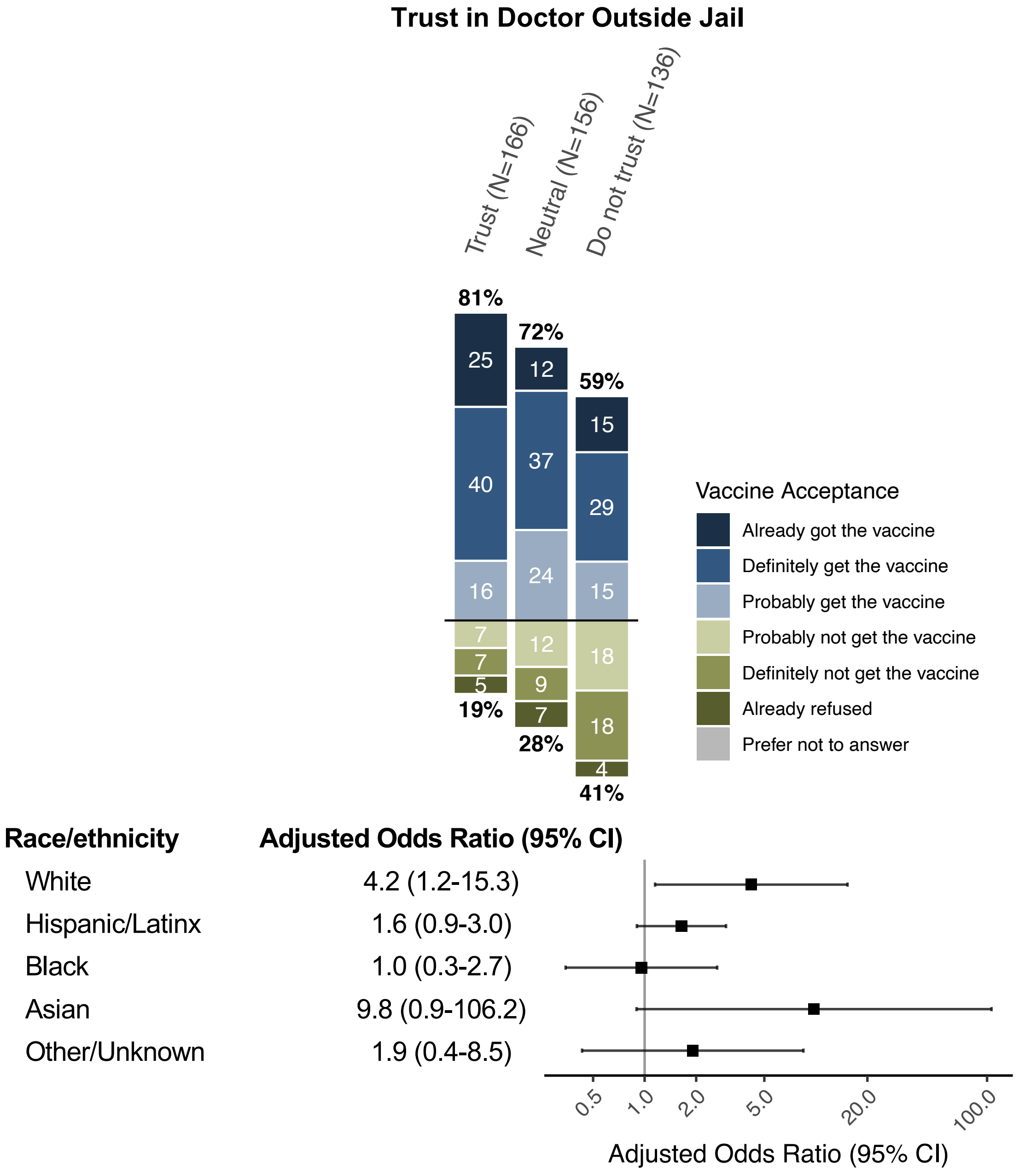

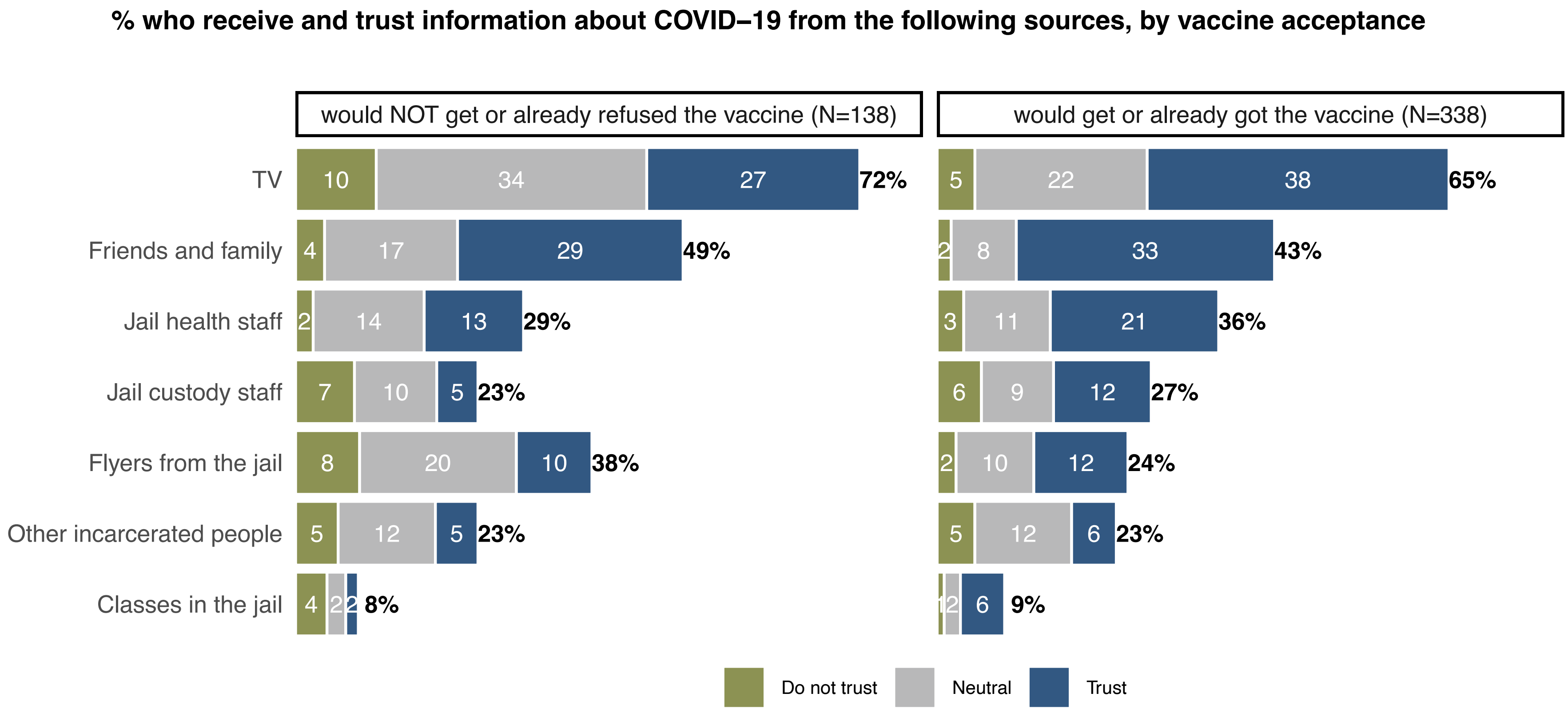
