## Supplementary material for "Factors associated with COVID-19 vaccine acceptance and hesitancy among residents of Northern California jails": S1 Survey

**The following questions will ask about your attitudes toward vaccines and medical care. Vaccination is NOT a part of this study. Like the rest of this questionnaire, your answers will be confidential and will not affect your housing, sentence, parole, medical care, or participation in this study.**

| | |
|---|---|
| Have you been vaccinated for COVID 19? | ◯ Yes<br>◯ No |
| When were you vaccinated? | __________________________<br>( (the first dose of COVID-19 vaccine)) |
| If a vaccine to prevent COVID-19 were available today, I would choose to: | ◯ Definitely get the vaccine<br>◯ Probably get the vaccine<br>◯ Probably not get the vaccine<br>◯ Definitely not get the vaccine<br>◯ Prefer not to answer |
| Consider another possible situation: a COVID-19 vaccine has been approved by the FDA, but supply is limited. If it were only available to high risk groups including healthcare workers and people in jails/prisons, I would choose to: | ◯ Definitely get the vaccine<br>◯ Probably get the vaccine<br>◯ Probably not get the vaccine<br>◯ Definitely not get the vaccine<br>◯ Prefer not to answer |
| If you probably or definitely would not get a vaccine to prevent COVID-19, please select the reasons you would not below: | ☐ Concern about side effects<br>☐ Do not think I need it<br>☐ It would cost too much<br>☐ Want to know more about how well it works<br>☐ Other<br>☐ Prefer not to answer |
| Other: Please Specify | __________________________ |
| Is "concern about side effects" a major or minor reason you would not get the vaccine? | ◯ Major reason<br>◯ Minor reason<br>◯ Prefer not to answer |
| Is "I do not think I need it" a major or minor reason you would not get the vaccine? | ◯ Major reason<br>◯ Minor reason<br>◯ Prefer not to answer |
| Is "It would cost too much" a major or minor reason you would not get the vaccine? | ◯ Major reason<br>◯ Minor reason<br>◯ Prefer not to answer |
| Is "Want to know more about how well it works" a major or minor reason you would not get the vaccine? | ◯ Major reason<br>◯ Minor reason<br>◯ Prefer not to answer |
| Which of the following factors would make you less likely to get the vaccine? | ☐ I would have to pay out of pocket to get it<br>☐ If many people experienced minor side effects<br>☐ If the vaccine was effective about 60% of the time<br>☐ If I needed to get the vaccine again every year<br>☐ Prefer not to answer |
| "I would have to pay out of pocket to get a vaccine" would make me: | ◯ A LOT less likely to get a vaccine<br>◯ A LITTLE less likely to get a vaccine<br>◯ Would not make a difference in if I got a vaccine<br>◯ Prefer not to answer |
| "If many people experienced minor side effects" would make me: | ◯ A LOT less likely to get a vaccine<br>◯ A LITTLE less likely to get a vaccine<br>◯ Would not make a difference in if I got a vaccine<br>◯ Prefer not to answer |

| | |
|---|---|
| "If the vaccine was effective about 60% of the time" would make me: | ◯ A LOT less likely to get a vaccine<br>◯ A LITTLE less likely to get a vaccine<br>◯ Would not make a difference in if I got a vaccine<br>◯ Prefer not to answer |
| "If I needed to get the vaccine again every year" would make me: | ◯ A LOT less likely to get a vaccine<br>◯ A LITTLE less likely to get a vaccine<br>◯ Would not make a difference in if I got a vaccine<br>◯ Prefer not to answer |
| From where do you receive information about COVID-19? | ☐ Correctional officers/custody staff<br>☐ Correctional health staff (i.e. doctors and nurses)<br>☐ Educational programming in the jail (i.e. classes)<br>☐ Friends and family<br>☐ Informational flyers from the jail<br>☐ Other incarcerated people<br>☐ TV news and other sources of news<br>☐ Other<br>☐ Prefer not to answer |
| I trust information about COVID-19 that I receive from correctional officers/custody staff | ◯ Strongly disagree<br>◯ Disagree<br>◯ Neutral<br>◯ Agree<br>◯ Strongly agree<br>◯ Prefer not to answer |
| I trust information about COVID-19 that I receive from correctional health staff (i.e. doctors and nurses) | ◯ Strongly disagree<br>◯ Disagree<br>◯ Neutral<br>◯ Agree<br>◯ Strongly agree<br>◯ Prefer not to answer |
| I trust information about COVID-19 that I receive from educational programming in the jail (i.e. classes) | ◯ Strongly disagree<br>◯ Disagree<br>◯ Neutral<br>◯ Agree<br>◯ Strongly agree<br>◯ Prefer not to answer |
| I trust information about COVID-19 that I receive from friends and family | ◯ Strongly disagree<br>◯ Disagree<br>◯ Neutral<br>◯ Agree<br>◯ Strongly agree<br>◯ Prefer not to answer |
| I trust information about COVID-19 that I receive from informational flyers from the jail | ◯ Strongly disagree<br>◯ Disagree<br>◯ Neutral<br>◯ Agree<br>◯ Strongly agree<br>◯ Prefer not to answer |
| I trust information about COVID-19 that I receive from other incarcerated people | ◯ Strongly disagree<br>◯ Disagree<br>◯ Neutral<br>◯ Agree<br>◯ Strongly agree<br>◯ Prefer not to answer |
| I trust information about COVID-19 that I receive from TV news and other sources of news | ◯ Strongly disagree<br>◯ Disagree<br>◯ Neutral<br>◯ Agree<br>◯ Strongly agree<br>◯ Prefer not to answer |

**As you read each of the following statements, please think about the medical care that you have received outside of jails/prisons. If you have not received any medical care recently, select the answer based on what you would expect if you had to seek medical care outside of jails/prisons today.**

| | |
|---|---|
| I trust my doctor's judgments about my medical care. | ◯ Strongly Disagree<br>◯ Disagree<br>◯ Neutral<br>◯ Agree<br>◯ Strongly Agree<br>◯ Prefer not to answer |
| I trust my doctor to put my medical needs above all other considerations when treating my medical problems. | ◯ Strongly Disagree<br>◯ Disagree<br>◯ Neutral<br>◯ Agree<br>◯ Strongly Agree<br>◯ Prefer not to answer |
| I believe that my doctor takes my health concerns seriously. | ◯ Strongly Disagree<br>◯ Disagree<br>◯ Neutral<br>◯ Agree<br>◯ Strongly Agree<br>◯ Prefer not to answer |
| While outside of jail/prison, I worry that I will be denied the treatment or services I need. | ◯ Strongly Disagree<br>◯ Disagree<br>◯ Neutral<br>◯ Agree<br>◯ Strongly Agree<br>◯ Prefer not to answer |
| Generally speaking, would you say that most people can be trusted or that you can't be too careful in dealing with people? | ◯ Can trust<br>◯ Cannot trust<br>◯ Depends<br>◯ Prefer not to answer |

**As you read each of the following statements, please think about the medical care that you have received in jails/prisons. If you have not received any medical care recently, select the answer based on what you would expect if you had to seek medical care in a jail or prison today.**

| | |
|---|---|
| I trust the jail doctors' and nurses' judgments about my medical care. | ◯ Strongly Disagree<br>◯ Disagree<br>◯ Neutral<br>◯ Agree<br>◯ Strongly Agree<br>◯ Prefer not to answer |
| I trust the jail doctors and nurses to put my medical needs above all other considerations when treating my medical problems. | ◯ Strongly Disagree<br>◯ Disagree<br>◯ Neutral<br>◯ Agree<br>◯ Strongly Agree<br>◯ Prefer not to answer |
| I believe that the jail doctors and nurses take my health concerns seriously. | ◯ Strongly Disagree<br>◯ Disagree<br>◯ Neutral<br>◯ Agree<br>◯ Strongly Agree<br>◯ Prefer not to answer |
| While in jail/prison, I worry that I will be denied the treatment or services I need. | ◯ Strongly Disagree<br>◯ Disagree<br>◯ Neutral<br>◯ Agree<br>◯ Strongly Agree<br>◯ Prefer not to answer |
