## Supplementary material for "Factors associated with COVID-19 vaccine acceptance and hesitancy among residents of Northern California jails": S2 Text

**Methods for linkage of survey data with vaccine uptake data.**

We accessed survey respondents’ vaccine uptake data through June 30, 2021 from their electronic health record (EHR) in Santa Clara County (SCC) and from Correctional Health Services in San Mateo County (SMC). We linked survey and vaccine uptake data by performing fuzzy string matching (*fuzzyjoin* in R) between self-reported names from the survey and names in the EHR or vaccination record.

In SCC, 355 participants were matched with the EHR, of which 215 were vaccinated, 104 had refused, and 36 had no vaccination data. In SMC, 72 participants were vaccinated according to Correctional Health Services’ vaccination record, and 77 participants were missing from the record. Because the SMC record only included vaccinations, not refusals, we accessed additional data on release dates for these 77 participants to determine whether they had refused the vaccine or were lost to follow-up due to release from custody. Of these 77 participants, 19 were released prior to 1/28/21 (when vaccinations began for medically vulnerable people in SMC jails) and were deemed to be lost to follow-up. 27 participants were deemed to have refused the vaccine for having been 1) in custody throughout the first period of mass vaccination (3/19/21-3/27/21) and/or 2) incarcerated for 14 days or more after this first period. We could not determine either loss to follow-up or refusal for the remaining 31 participants who were released between 1/28/21 and 3/27/21 and/or were in custody for less than 14 days.

For respondents whose demographic information (age, gender, race/ethnicity) or incarceration start date were missing, we also accessed this information in their EHR (SCC) or custody record (SMC). Five of 360 participants in SCC and one of 149 participants in SMC could not be matched with the EHR or custody record, respectively, likely due to errors in self-reported name.
